# Data-driven Optimized Control of the COVID-19 Epidemics

**DOI:** 10.1101/2020.08.27.20183574

**Authors:** Afroza Shirin, Yen Ting Lin, Francesco Sorrentino

## Abstract

Optimizing the impact on the economy of control strategies aiming at containing the spread of COVID-19 is a critical challenge. We use daily new case counts of COVID-19 patients reported by local health administrations from different Metropolitan Statistical Areas (MSAs) within the US to parametrize a model that well describes the propagation of the disease in each area. We then introduce a time-varying control input that represents the level of social distancing imposed on the population of a given area and solve an optimal control problem with the goal of minimizing the impact of social distancing on the economy in the presence of relevant constraints, such as a desired level of suppression for the epidemics at a terminal time. We find that with the exception of the initial time and of the final time, the optimal control input is well approximated by a constant, specific to each area, which contrasts with the implemented system of reopening ‘in phases’. For all the areas considered, this optimal level corresponds to stricter social distancing than the level estimated from data. Proper selection of the time period for application of the control action optimally is important: depending on the particular MSA this period should be either short or long or intermediate. We also consider the case that the transmissibility increases in time (due e.g. to increasingly colder weather), for which we find that the optimal control solution yields progressively stricter measures of social distancing. We finally compute the optimal control solution for a model modified to incorporate the effects of vaccinations on the population and we see that depending on a number of factors, social distancing measures could be optimally reduced during the period over which vaccines are administered to the population.

## Introduction

The fast propagation of the COVID-19 pandemic has attracted unprecedented attention from both the public and the scientific community. This has resulted in much research and funding getting redirected towards COVID-19 and stimulated strong research collaboration between countries [34]. Due to the high fatality rate of SARS-CoV-2 [37], governments throughout the world have adopted measures such as lock-down, stay-at-home, and shelter-in-place, which in turn have led to substantial economic losses, see e.g., [24]. In many countries control interventions have been articulated in phases, usually phase 1 to phase 3, with higher phases corresponding to progressively lower restrictions [14]. A fundamental challenge is to balance the need to suppress the spread of COVID-19 and the need to contain the economic impact of measures aiming at limiting the spread of the disease. In this manuscript, we apply optimal control theory on a mathematical model for the propagation of the epidemics, parametrized by real-world data describing different regions, and compute control strategies which are optimal for each region from an economic standpoint.

A number of papers and reports have focused on both modeling and controlling the pandemic. Flaxman et al. [12] looked at the effect of non-pharmaceutical interventions including school closures, banning of mass gathering, social distancing, etc. on the reproductive number *R*_0_ of COVID-19. Sanche et al. [40] used a mathematical model with data on individual cases, real-time human travel, and infections, as well as estimated epidemiology parameters to compute *R*_0_ and found that it is higher than initially estimated. Chang et al. [9] adopted an agent-based model to determine the efficacy of several intervention strategies on the spread of COVID-19 in Australia. Anderson et al. [6] performed the Bayesian analysis to estimate the impact of social distancing on number of reported cases and hospitalizations in British Columbia and found that when 78% participation in social distancing has been accomplished the cases would decrease; it also noted that if the participation were below 45%, an exponential growth would restart. Morris et al. [27] explored COVID-19 intervention methods which are robust to implementation errors and found that these methods in conjunction with optimal time-limited methods derived from the standard SIR model can be used to mitigate the spread of the virus. Another study analyzed an open-loop optimal control policy updated weekly using real-world feedback, and found that this method is effective in reducing fatalities even if some measurements are inaccurate [18]. A study published in March 2020 estimated the ICU occupancy and ventilator use from a statistical model under the conditions of social distancing and found that the demand for hospital beds and ventilators will exceed the current supply [11]. Another study explored optimal policies for decreasing economic cost and mortality rates from a multi-risk SIR model and found that strict lock-down policies which specifically target the elderly population were most effective in minimizing deaths and economic losses [5]. Previous work has not computed optimal controls for data driven models. On the one hand, Refs. [9], [27], [18], [5] explicitly compute optimal control strategies, but their models were stylized and not parametrized/calibrated by data; On the other hand, Refs. [12] [40], [6], [11] used data to parametrize the models, but the models do not have controllers which can be used to infer optimal control strategies.

In what follows, we first construct a mathematical model, which is parametrized by historical and regional daily new case report. After parametrization, the data-driven model is capable to reproduce the regional progression of the COVID-19 epidemics up to the present. Then, we apply optimal control theory to the parametrized model to compute an optimal control strategy over the course of a pre-determined period into the future to suppress the epidemics to a desired level, while minimizing economic costs. This type of approach is suitable for long-term planning (i.e., over the course of several months) as opposed to short-term planning, which can be difficult to implement by the government and by businesses.

In most countries, distribution of the vaccine to the population is under way and will continue for most of 2021, which points out the need for planning interventions to contain the epidemics for several months while only a limited part of the population has received a vaccine. Thus our proposed workflow aims to bridge the gap until the time *T*_*vac*_ when an effective vaccine is massively manufactured, and administered to the majority of the population. Another temporal consideration regards the time *T*_*herd*_ at which a population achieves herd immunity in the absence of a vaccine and without overflowing the medical facilities. While herd immunity from COVID-19 is the subject of much ongoing discussion [36], in the Methods we provide a rough estimate of *T*_*herd*_ from available data. In this paper we proceed under the assumption that the inference period *T*_*inf*_ and the control horizon *T*_*cont*_ are such that *T*_*inf*_ + *T*_*cont*_ *< T*_*vac*_ and *T*_*inf*_ + *T*_*cont*_ *< T*_*herd*_.

We set out the analysis by first introducing a compartmental model which describes key features of the COVID-19 epidemics. The whole population is divided into the following compartments. The susceptible population compartment (S) includes the people who can contract the pathogen SARS-CoV-2. The exposed population (E) are those who have been infected but have not progressed long enough into the disease to transmit it to susceptible people. Those who can transmit the disease (‘carriers’) are divided into the asymptomatic group (A) who do not show symptoms and the infected symptomatic groups (I) who show symptoms. Both the symptomatic and asymptomatic groups can transmit the disease, but with different infectiousness—the asymptomatic people are less infectious. The infected symptomatic population *I* is divided into three sub-compartments. The first sub-compartment *I*_*sq*_ includes those who just self quarantine and do not get tested. The second sub-compartment *I*_*tp*_ includes those who get tested, result positive, and get quarantined. In contrast to the previous two sub-compartments, the third sub-compartment *I*_*s*_ includes the rest of the symptomatic population, who do not change their behavior until they recover (or decease). Those who were tested positive and those who decided to self-quarantine are moved into a quarantined compartment (Q), and stop interacting with other populations. The removed compartment (R) includes those who are completely recovered from the disease and have acquired immunity, and those who have died because of the disease. Both groups are not susceptible to reinfection and are removed from the system.

Mathematically, the time evolution of the population density—defined as the compartmental population normalized by the total regional population—of each compartment is governed by the following set of coupled ordinary differential equations,

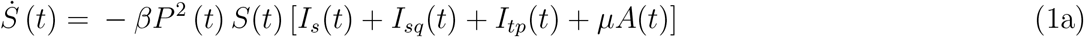

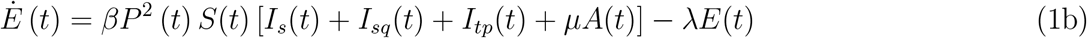

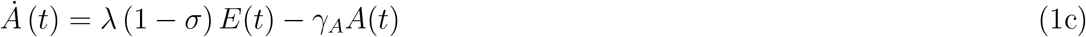

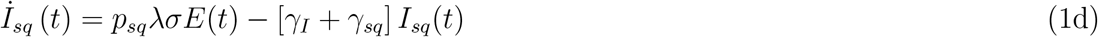

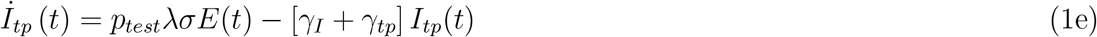

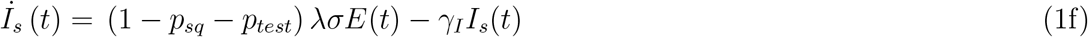

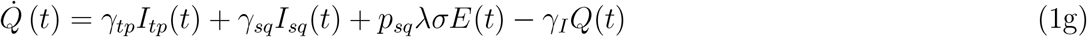

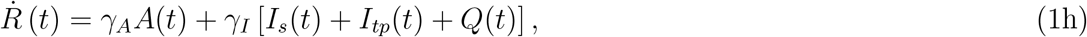

where *β* is the transmissibility, *λ* is the transition rate from the exposed compartment to either the asymptomatic or symptomatic compartments, *γ*_*I*_ is the transition rate from the infected compartment to the recovered compartment, *γ*_*A*_ is the transition rate from the asymptomatic compartment to the recovered compartment, *µ* is the relative infectiousness of asymptomatic individuals (compared to symptomatic individuals), *σ* is the fraction of exposed people who transition into the symptomatic compartments, *p*_*sq*_ is the fraction of symptomatic people who will self-quarantine, *γ*_*sq*_ is the transition rate from from the *I*_*sq*_ compartment into the quarantined compartment, *γ*_*tp*_ is the transition rate from the *I*_*tp*_ compartment into the quarantined compartment, and *p*_*test*_ is the fraction of infected who get tested (but only a fraction of them will be able to get a test and be confirmed as a positive, *I*_*tp*_). The model assumes that testing resources are not scarce, i.e., availability of a testing kit to every person in *I*_*tp*_; we also assume positive people are identified as such with 100% accuracy. The case with limited testing resources is discussed in the SI. Realistically, the time scale associated with *γ*_*tp*_ can be several days, so we assume *γ*_*tp*_ = 0.5 (2 days). We model social distancing by the control variable 0 ≤ *P* (*t*) ≤ 1, which measures the reduction of contact probabilities between the susceptible and the infectious populations (which include both A and I). The model is structurally similar to the models analyzed in Refs. [6, 23], but simplified to allow for efficient calculations of optimal control solutions. Figure 1 illustrates a schematic diagram of the model.

**Figure 1:**
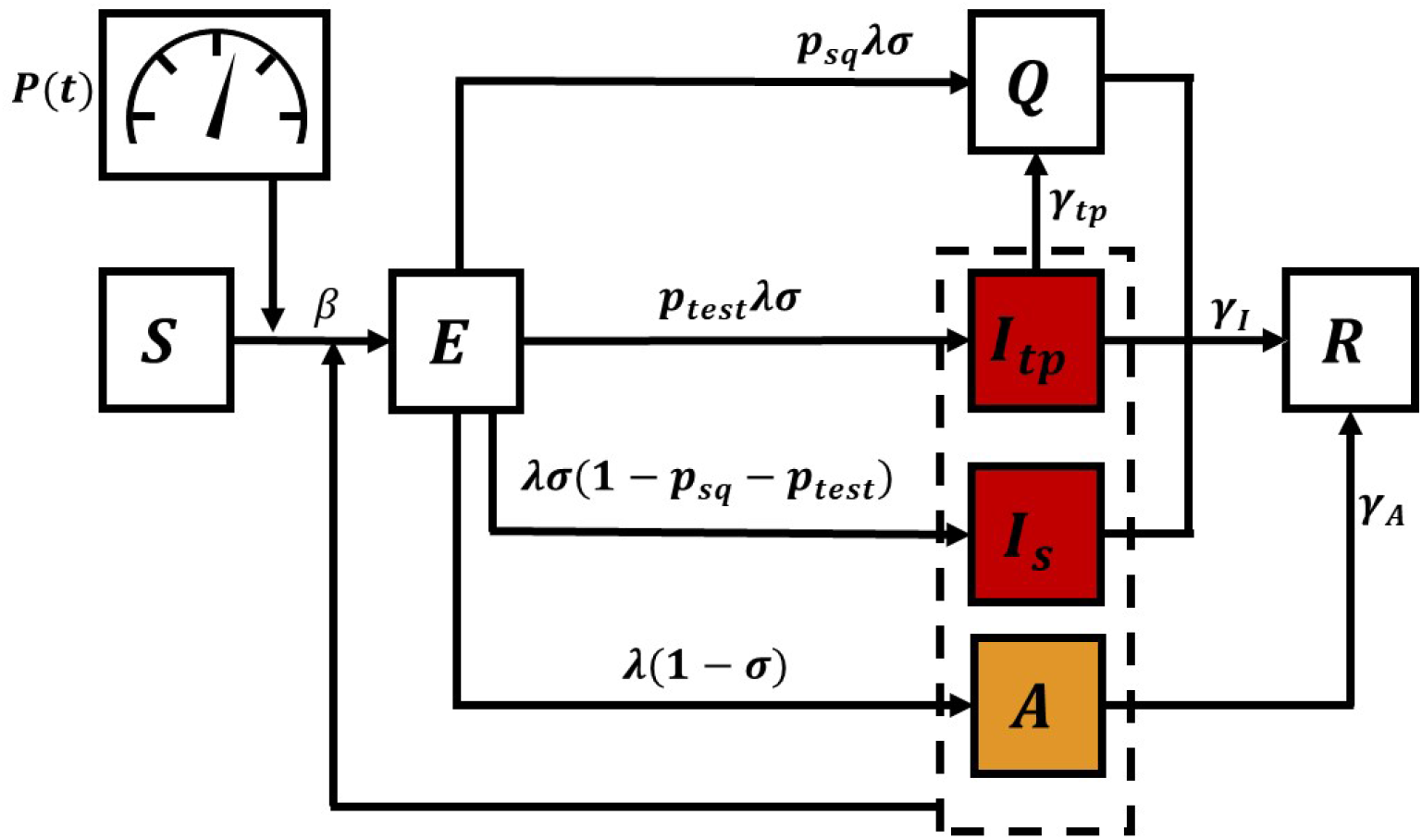
Compartmental model corresponding to Eq. 2. The transition from the *S* (susceptible) compartment to the *E* (exposed) compartment is affected by the population densities *S*(*t*), *I*_*tp*_(*t*), *I*_*s*_(*t*), *A*(*t*) and by the time-varying control input *P* (*t*).

In order to reduce the dimensionality of the dynamical system, we introduce the following simplification: we treat *γ*_*sq*_ as a very large number; for *γ*_*sq*_ → ∞, we assume that those becoming symptomatic immediately transition into the *Q* compartment, yielding the following reduced-order model:

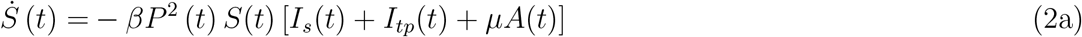

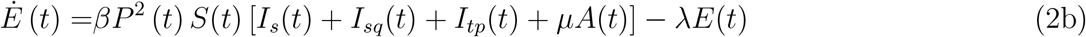

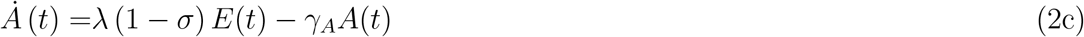

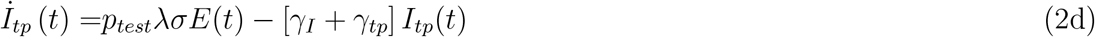

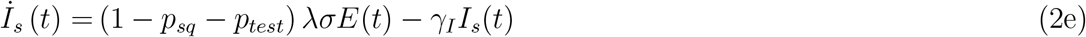

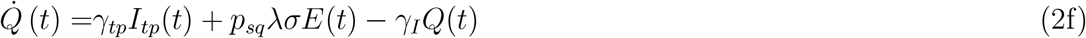

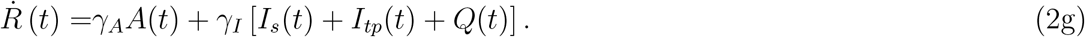

The schematic diagram which fits the above model in Eq. (2) is shown in the Supplementary Note 1 1.

To bridge the model and the data, we also integrate an auxiliary variable *C*_*I*_ which counts the cumulative confirmed cases and evolves according to

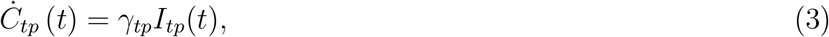

We will fit Δ*C*_*tp*_(*t*) := *C*_*tp*_(*t* + 1) −*C*_*tp*_(*t*) to the new case counts reported on each day, detailed in the section on parametrization below.

From an economic point of view, the measures of social distancing *P* (*t*) and quarantining *Q*(*t*) have very elevated costs for society. For example, the US real gross domestic product (GDP) dropped by roughly one third from year to year in the second quarter of 2020 [24], due mainly to the COVID-19 pandemic, see e.g., [28].

We thus formulate the following optimal control problem,

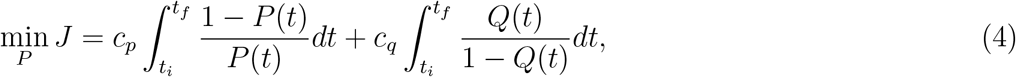

*c*_*p*_, *c*_*q*_ ≥ 0, subject to Eq. (2), the initial conditions, the terminal constraint

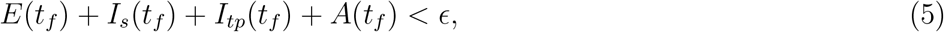

and the following path constraint

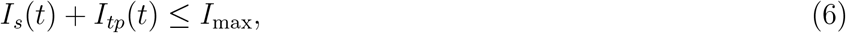

where *I*_max_ is an upper limit on the number of infected people that can receive proper treatment in the hospitals. *t*_*i*_ is the time at which we perform the inference and start optimizing the control action, *t*_*f*_ is the final time of the optimization, the previously defined control horizon *T*_*cont*_ = (*t*_*f*_ −*t*_*i*_).

The objective function (4) takes into account an economic cost for social distancing with an appropriate coefficient *c*_*p*_ *>* 0 and an economic cost for quarantining with an appropriate coefficient *c*_*q*_ *>* 0. We set the cost associated with social distancing per unit time to be modeled by (1 − *P* (*t*)) */P* (*t*). By choosing this functional form, the cost is linearly dependent on the scale of the reduction (1 − *P* (*t*)) when 1 −*P* (*t*) « 1, and nonlinearly diverges near total shut-down (*P* (*t*) « 1). The economic cost increases with 1 − *P* (*t*) to indicate that stricter measures of social distancing affect more and more ‘essential’ workers, and so increasingly larger parts of the economy. For example, the cost of limiting large gatherings of people like concerts or sport events is lower than the cost of limiting customers’ access to stores and restaurants. Similarly, imposing quarantine requires resources that are linearly proportional to the quarantined population when *Q* is small, and diverge nonlinearly when the quarantined population is close to the entire population *Q* = 1. We model such a cost by the functional form *Q*(*t*)*/* (1 −*Q*(*t*)).

Both social distancing and quarantining have a cost associated with the lack of economic return generated by limiting person-to-person interactions. It is reasonable to assume *c*_*q*_ ≥ *c*_*p*_, as strict quarantining requires supervision costs as well as costs due to lowered productivity [15] while social distancing only incurs costs due to lowered productivity [19].

We will also consider the alternative objective function,

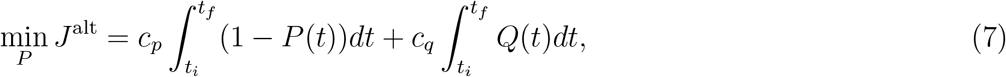

for which the integrand functions are linear in *P* (*t*) and *Q*(*t*). While the formulation (7) is mathematically simpler, it does not take into account the fact that stricter measures of social distancing may result in progressively larger economic losses. The alternative objective function (7) is here mainly introduced in order to compare the results to those obtained with (4).

The tunable parameter *ϵ* represents the desired suppression level for the epidemics at the final time *t*_*f*_. In general, selected values for *ϵ* may depend on a number of factors, such as the time horizon over which optimization is performed and the particular stage of the epidemics (initial exponential growth, intermediate growth, or plateau.) A possibility is to require complete eradication of the epidemics, which corresponds to setting *ϵ* = 1*/N*, where *N* is the number of individuals in the population. However, the high basic reproductive number of COVID-19 makes eradication unlikely; instead we set *ϵ* to a small number indicating suppression of the disease. The other constraint given by *I*_max_ represents the need to contain the infected population below a given threshold at all times (or ‘flatten the curve’.) In what follows, unless differently stated, we set the terminal constraint *ϵ* = 10^−5^, which corresponds to imposing that the number of infected people is below one person per 100000 population. This number is derived from the guidelines of European countries about re-opening, see for example [26], indicating that reopening occurred at about 2 × 10^−5^. Also, European countries have official guidelines for reimposing stricter lock-down measures, see: [13], which sets a critical population equal to 50*/*100000 = 5 × 10^−4^. The values of *I*_*max*_ are set regionally based on the capacity of the ICU beds in different metropolitan areas and are summarized in Table 3.

**Table 1:**
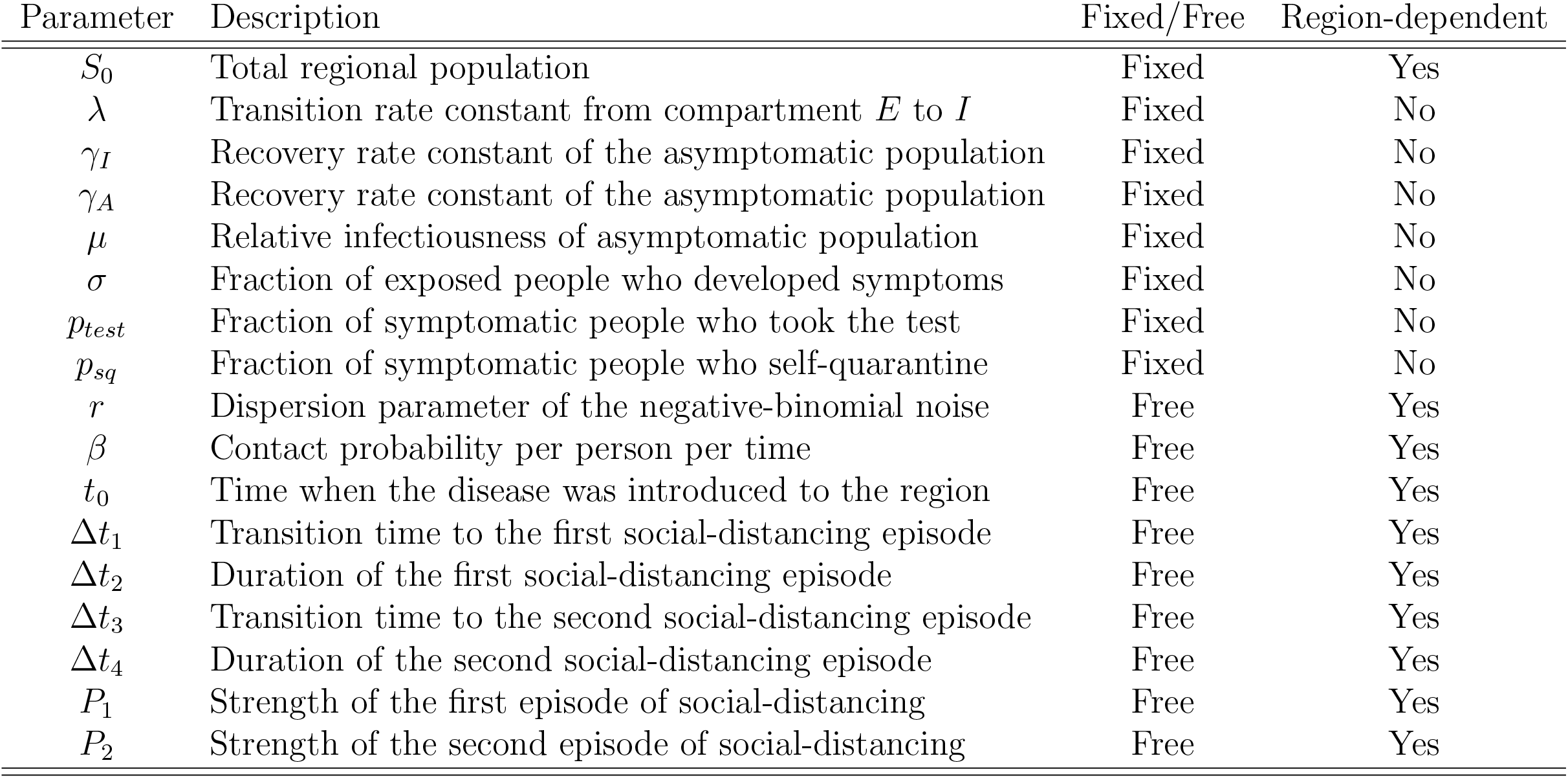
List of data-driven model parameters.

**Table 2:** Parameters by Metropolitan Statistical Area (MSA)

| Parameter | New York | Los Angeles | Houston | Seattle |
| --- | --- | --- | --- | --- |
| $S_0$ | 1.92E+07 | 1.321E+07 | 7.066E+06 | 3.979E0+6 |
| $\lambda$ | 3.333E-01 | 3.333E-01 | 3.333E-01 | 3.333E-01 |
| $\gamma_I$ | 1.200E-01 | 1.200E-01 | 1.200E-01 | 1.200E-01 |
| $\gamma_A$ | 2.60E-01 | 2.60E-01 | 2.60E-01 | 2.60E-01 |
| $\mu$ | 9.00E-01 | 9.00E-01 | 9.00E-01 | 9.00E-01 |
| $\sigma$ | 5.60E-01 | 5.60E-01 | 5.60E-01 | 5.60E-01 |
| $p_{test}$ | 2.50E-01 | 2.50E-01 | 2.50E-01 | 2.50E-01 |
| $p_{sq}$ | 4.00E-01 | 4.00E-01 | 4.00E-01 | 4.00E-01 |
| $r$ | 1.183E+01 | 1.100E+01 | 5.287 | 1.067E+01 |
| $\beta$ | 1.806 | 1.204 | 1.123 | 1.442 |
| $t_0$ | 2.911E+01 | 2.865E+01 | 3.195E+01 | 2.339E+01 |
| $\Delta t_1$ | 2.557E+01 | 3.237E+01 | 3.153E+01 | 1.388E+01 |
| $\Delta t_2$ | 2.148E+01 | 1.153E+01 | 1.633E+01 | 3.549E+01 |
| $\Delta t_3$ | 6.423E+01 | 7.171E+01 | 1.685E+01 | 6.614E+01 |
| $\Delta t_4$ | 8.800 | 4.289 | 4.658E+01 | 2.016 |
| $P_1$ | 3.693E-01 | 5.371E-01 | 4.845E-01 | 4.160E-01 |
| $P_2$ | 4.403E-01 | 5.968E-01 | 6.391E-01 | 5.555E-01 |

**Table 3:** *I*_max_ values for U.S. Cities

| City | $I_{\max}$ ($\rho = 2/3$) | $I_{\max}$ ($\rho = 1$) |
| --- | --- | --- |
| New York, NY | 0.88%(5704/648000) | 1.32% (8556/648000) |
| Los Angeles, CA | 0.64% (8241/1280124) | 0.97%(12361/1280124) |
| Chicago, IL | 1.00%(4094/410508) | 1.50%(6141/410508) |
| Dallas, TX | 0.86%(8109/939600) | 1.29%(12163/939600) |
| Houston, TX | 0.86%(8109/939600) | 1.29%(12163/939600) |
| Riverside, CA | 0.64% (8241/1280124) | 0.97%(12361/1280124) |
| Miami, FL | 0.98% (6829/695952) | 1.47%(10244/695952) |
| Philadelphia, PA | 1.047%(4342/414720) | 1.57%(6513/414720) |
| Atlanta, GA | 0.65%(2249/344088) | 0.98%(3374/344088) |
| Phoenix, AZ | 0.55%(1309/2358450) | 0.83%(1963/2358450) |
| Boston, MA | 0.4%(893/223330) | 0.60%(1340/223330) |
| San Francisco, CA | 0.64% (8241/1280124) | 0.97%(12361/1280124) |
| Seattle, WA | 0.46%(1143/246888) | 0.69%(1714/246888) |
| Indianapolis, IN | 0.83%(1811/218120) | 1.25%(2716/218120) |
| Detroit, MI | 0.71%(2315/32358) | 1.07%(3472/32358) |
| Baltimore, MD | 0.40%(893/223330) | 0.60%(1340/223330) |
| Denver, CO | 0.56%(1039/186590) | 0.84%(1559/186590) |
| Portland, OR | 0.56%(768/136660) | 0.84%(1152/136660) |
| Las Vegas, NV | 0.75%(752/99792) | 1.13%(1129/99792) |

### Parametrization of the model for different US metropolitan areas

We partitioned the model parameters into two sets: a set of fixed parameters and a set of inferred parameters which are estimated by the daily case counts reported by regional health administration and registered in the repository curated by the New York Times [3].

The fixed parameter sets includes *S*_0_, *λ, γ*_*I*_, *γ*_*A*_, *µ, σ, p*_*test*_, and *p*_*sq*_. *S*_0_ is the regional total population, and we used the US Census Bureau-estimated regional population of each of the Metropolitan Statistical Areas (MSAs) or ‘cities’, which are delineated by the US Office of Management and Budget [4]. Lauer et al. [21] estimated the median of the incubation period to be about 5.2 days, however, there is evidence that patients become infectious roughly two days before the onset of symptoms [29], which corresponds to approximately a three-day progression into *contagiousness*. We thus set a rough estimation for *λ* to be 1*/*3 (d). In a more complex model with multiple stages of the disease progression [23], one could account for the fact that a patient can be both pre-symptomatic and infectious. The coefficients *γ*_*I*_ and *γ*_*A*_, which are the transition rate to recovery of the symptomatic and asymptomatic populations, are estimated to be 0.12 (1/d) [47] and 0.26 (1/d) [39]. The relative infectiousness *µ* is set at 0.9 [45], and the fraction of the symptomatic population is set at 0.64 [2, 39]. The parametrization of *γ*_*I*_, *γ*_*A*_, *µ*, and *σ* are consistent with a more complex model which was used to perform daily forecasts of the disease spread [23]. We assume *p*_*test*_ = 0.25 and *p*_*sq*_ = 0.4, noting that these parameters were able to reproduce the infected population at the time of the inference (we estimated that about 15 to 20% of the total population infected in the New York City MSA on 07-Jul-2020 when we parametrize the model.)

We used the data from 21-Jan-2020 to 07-Jul-2020 to infer the inferred parameters, which corresponds to the previously defined inference period *T*_*inf*_. We assume that the social distance function *P* (*t*) before 07-Jul-2020 is piecewise-linear to avoid over-fitting (due to observation noise.) At the time when the analysis was performed, multiple MSAs had shown clear second-phase resurrection of the epidemics [23, 1]. We found that a two-phase piecewise linear function is sufficient to reproduce the data of each of the MSAs:

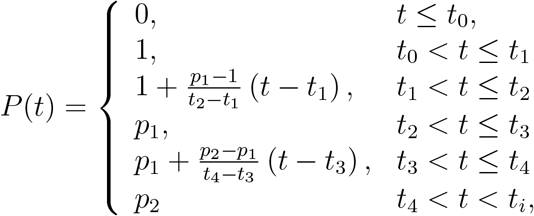

where *t*_0_ is the time when the disease was introduced into a specific MSA, the monotonic *t*_1_, *t*_2_, *t*_3_, and *t*_4_ (*t*_*j*_ ≤ *t*_*j*+1_) are the times when the social-distancing behavior changes, and *p*_1_ and *p*_2_ are the two social-distancing strengths. The two-phase model was selected by a model-selection procedure [23] and is deemed the most evident model structure (v. one- and three-phase). We define Δ*t*_*j*_ := *t*_*j*_ − *t*_*j*−1_ for *j* = 1, … 4. The time at which we perform the inference is *t*_4_, which is also the time after which optimization of the control action begins. The two-phase piecewise linear model is the minimal model that we found capable to reproduce the data, and can be validated by more rigorous model-selection procedure detailed in [23].

To fit the model by the noisy daily report new counts, we adopt a negative-binomial noise model. We brief the procedure here, noting that the procedure is similar to the inference method detailed in [23]. Given a set of parameters *θ* (a stylized notation of the set of the inferred model parameters), the model Eq. (2) predicts a deterministic trajectory of the *new* positive tested case on day *j*, Δ*I*_*tp*_(*j*; *θ*). This deterministic prediction is interpreted as the mean of a stochastic outcome, modeled by a negative binomial distribution NB(*r, p*_*j*_) where *r* and *p*_*j*_ are the parameter of the distribution, and *p*_*j*_ is constrained by the deterministic model prediction

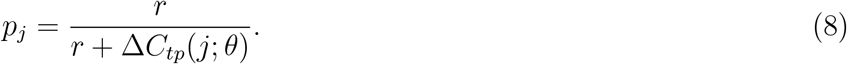

Here, the *r* is the *dispersion* parameter which describes how disperse the noise distribution is; in the limit *r* → ∞, the negative binomial statistics converges to Poissonian, and in the limit *r* → 0 the distribution looks closer to an exponential. The negative binomial noise model is phenomenological: it has the capability to capture a wide variety of single-modal distributions. With the negative binomial noise model, the likelihood function given a set of *N* daily reported new case counts 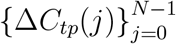 can be formulated [23]:

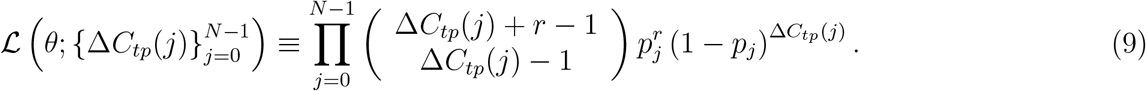

In summary, the inferred parameters *θ* include the regional-specific disease transmissibility *β*, onset of the disease spread *t*_0_, behavioral switching times *t*_1_, *t*_2_, *t*_3_, *t*_4_, the strengths of two social-distancing episodes *p*_1_, *p*_2_, and a dispersion parameter *r* of the negative binomial noise model. These parameters were inferred by the daily case reports from 21-Jan-2020 to 07-Jul-2020. With the formulated likelihood function (9), we used the standard Markov chain Monte Carlo procedure (MCMC) with an adaptive sampler ([7], detailed in [23]) to estimate the maximum likelihood estimator of the parameters *θ* for each of the interested MSA’s. Figure 2 shows excellent agreement between the daily new case counts reported by local health administrations (plus signs) and the daily new cases obtained by integrating Eq. (2) after parametrization of the model (solid line.) Table 1 is a list of data-driven model parameters, with indication of whether they are free or fixed and of whether they are region-dependent. Table 2 summarizes the set of model parameters that were estimated for each MSA.

**Figure 2:**
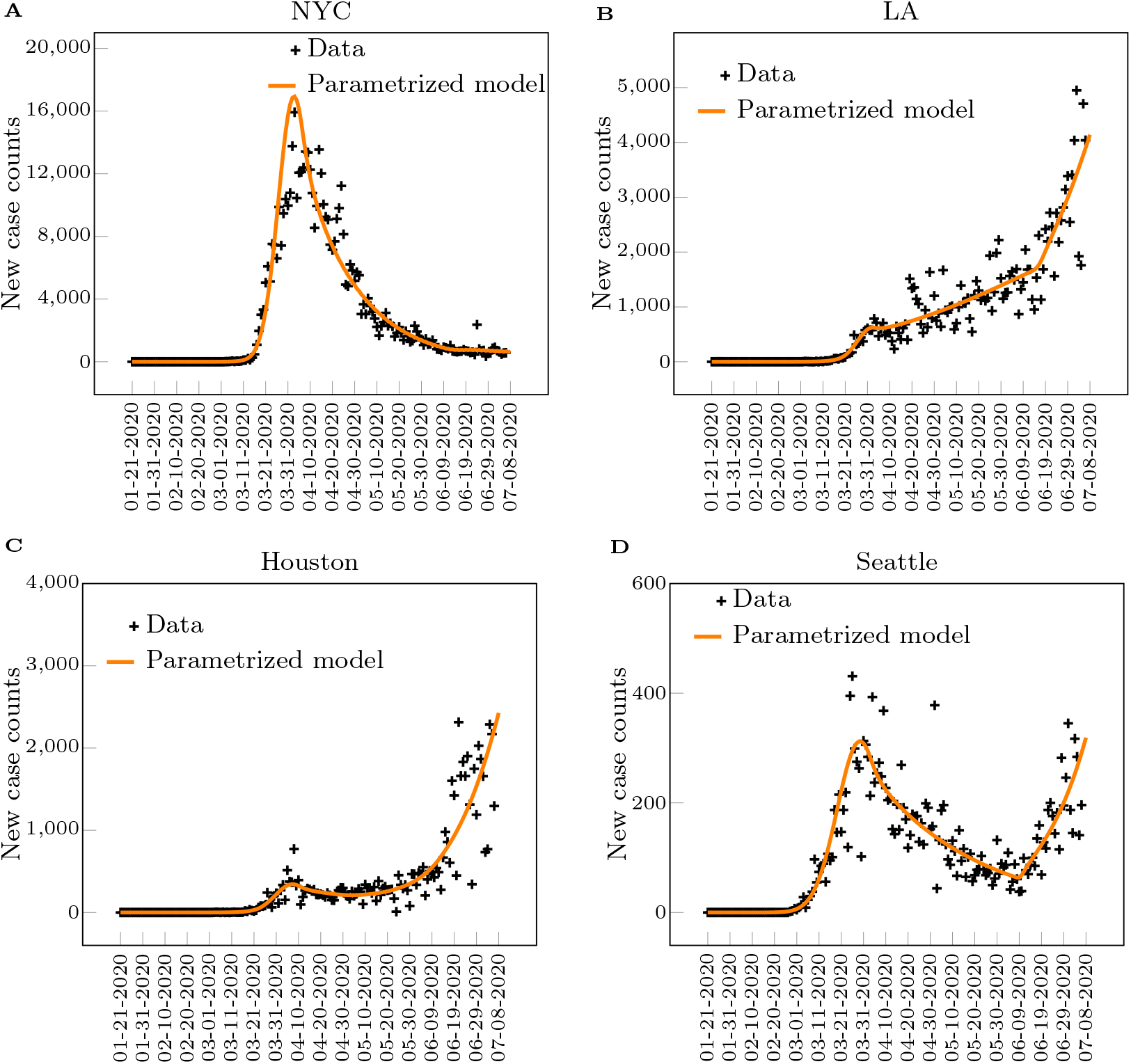
New case counts from January 21, 2020 to July 8, 2020 in four Metropolitan Statistical Areas within the US. Plus signs are daily new case counts reported by local health administrations. The solid line is the daily new cases obtained by integrating Eq. (2) after parametrization of the model, which was deemed to be sufficient to reproduce the trends in new daily case report [23]

### Results: Optimal Control Solutions

Following previous work by the authors [43, 42], we use pseudo-spectral optimal control (see Supplementary Note 7) to compute solutions for the problem formulated in Eqs. (2),(4),(5),(6). We set *t*_*i*_ = 169 (days) and *t*_*f*_ = 259, corresponding to a ninety day control horizon, (*t*_*f*_ − *t*_*i*_) = 90. We wish to emphasize that our choice of *t*_*i*_ is somehow arbitrary and coincides with the day when we initially computed the optimal control solutions. However, our procedure is pretty general and can be replicated for many possible choices of *t*_*i*_. We parametrize the solutions in *I*_max_, *E* and *c*_*q*_, after setting without loss of generality *c*_*p*_ = 1.

We focus on four different US metropolitan statistical areas: New York City (NYC), Los Angeles (LA), Houston, and Seattle. NYC is the largest US metropolitan area; it emerged as the main early hotspot of the epidemics in the US in March and April, but since early June has achieved stable control of the epidemics. LA is the second largest metropolitan area in the US, it has seen a steady rise in the number of cases by the time when we performed the inference. Houston has seen a rapid increase of the cases in June and July. Seattle was also a very early hotspot, which has seen a decrease in the number of cases in April and May, followed by an increase in June and July. We chose these four MSA’s to cover a wide range of different dynamics before the time of the inference.

The solution of the optimal control problem is the function of time *P* *(*t*) that minimizes the objective function (4) subject to the constraints (2), (5), and (6). Different from the observation period *t* ≤ *t*_*i*_, for which we set *P* (*t*) to be a piecewise linear function, in the optimization period *t* ∈ [*t*_*i*_, *t*_*f*_] we let *P* (*t*) be an arbitrary function of *t*, for the purpose of computing the optimal control solution.The optimal control solutions that we obtain for different MSAs are shown in Figs. 3. These solutions are robust to parameter variations (such as different values of the coefficient *c*_*q*_, see the Supplementary Note 2) and also qualitatively consistent for different MSA’s. Robustness is also found with respect to the choice of the constraint *I*_*max*_, see the Supplementary Note 2. Typically we first observe a quick drop in *P* *(*t*) (initial tightening), followed by a long nearly constant trend at 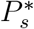 (steady social distancing), and by another drop near the final time (final stranglehold). We remark that 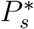 corresponds to the level of reduction needed to suppress the time-varying reproduction number *R*_*t*_ ≈ 1, and the cost per day associated with this level of reduction is 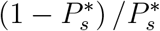. It follows that for each MSA, there is an almost constant value of 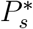 which well describes the optimal solution, except for the initial time and the final time. The value of 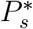 appears to be approximately the same for LA, Houston, and Seattle, while NYC has typically a slighter higher value of 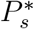. This also implies that the optimal cost function *J* * is lower for NYC than for the other cities. In all the four metropolitan areas, 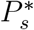 is lower than the ‘current’ value of *P* (*t*) estimated from data, as can be seen from the initial dip in *P* *(*t*). NYC has the smallest initial drop, indicating that the control action at the time at which we performed the inference is the closest to optimal, followed by (in the order) Seattle, LA, and Houston. For each metropolitan area, *I*_max_ was estimated from available data about ICU beds for different US states, as shown in Table 3 in the Methods.

**Figure 3:**
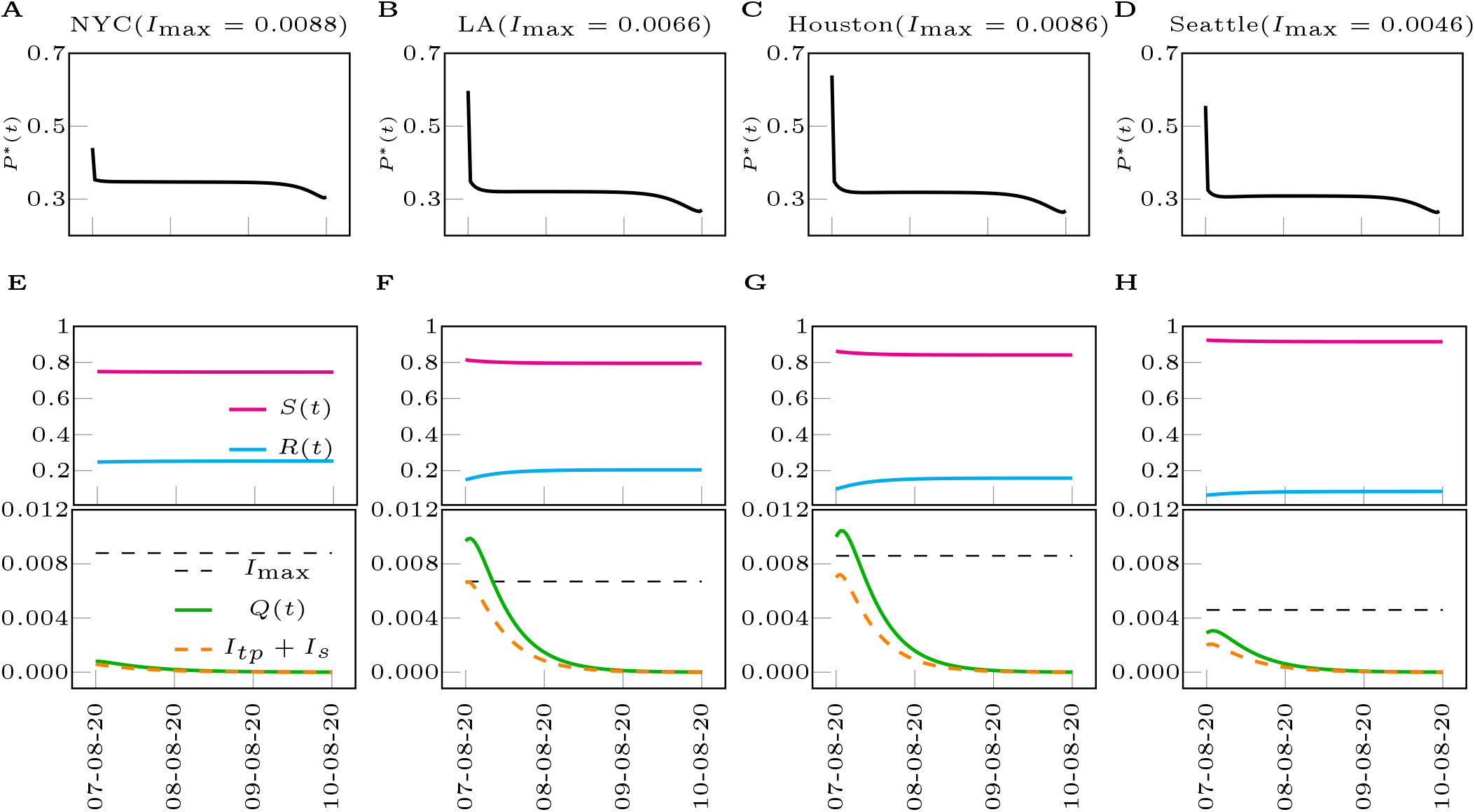
Optimal control solutions for different Metropolitan Statistical Areas within the US. (A-D) Optimal control inputs for the metropolitan cities of NYC, LA, Houston, Seattle (from left to right). (E-H) Time evolution of the states subject to the optimal control inputs. *I*_max_ are chosen as the minimum of the range in Table 3 (*ρ* = 2*/*3) and *c*_*p*_ = *c*_*q*_ = 1. The legends in (F-H) are same as the legend in (E).

The most important parameter of the optimal control problem is the terminal suppression constraint *ϵ*, which describes the level at which one is trying to suppress the epidemic. The lower is *ϵ*, the closer the goal is to eradication of the disease. The optimal value attained by the objective function *J* * versus *ϵ* is shown in Figs. 4(A) and (B). It should be noted that for each value of *ϵ, J* * shown in Fig. 4 is the lowest possible cost that can be attained. For all the US cities considered, this lowest possible cost increases dramatically as *ϵ* is reduced, which exemplifies the dilemma between saving human lives and saving the economy. Again, here we are assuming that the measures of social distancing are optimally implemented, while the cost would be higher in case of non-optimal implementation (discussed in the Supplementary Note 7). For large values of the suppression constraint *ϵ*, in all four cities considered, a different type of solution characterized by low cost emerges, which is discussed in more detail in what follows. Qualitatively similar results were obtained when the control input was chosen that minimizes the alternative objective function (7). A complete study of the effects of varying the suppression constraint parameter *ϵ* can be found in the Supplementary Note 3. There are deeper implications behind Figs. 4 (A) and (B), namely setting a larger value of *ϵ* corresponds to delaying the economic cost in time, rather than removing it.

**Figure 4:**
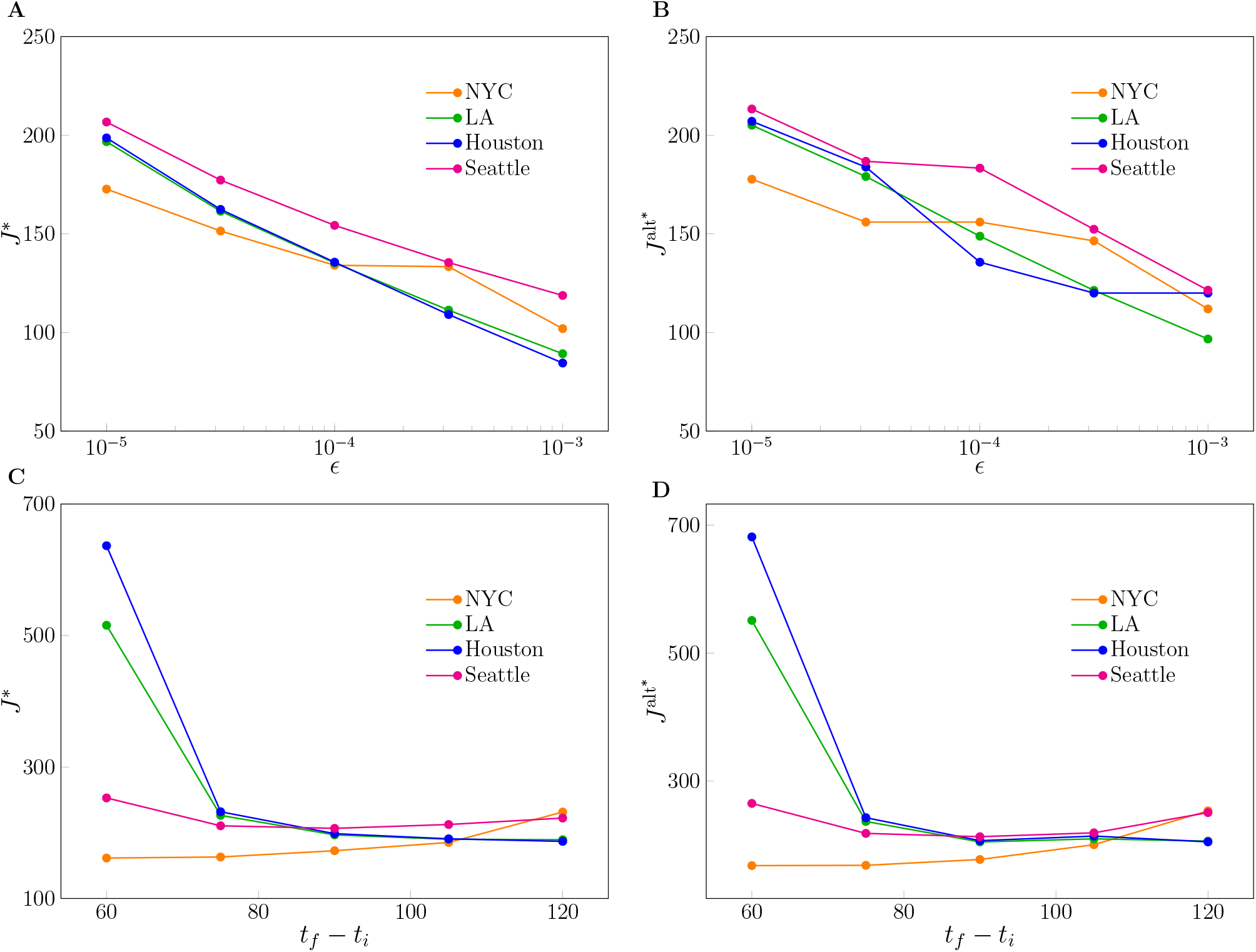
Effects of varying the terminal suppression constraint *ϵ* and the control horizon *t*_*f*_ − *t*_*i*_. (A) The optimal cost *J* * obtained as the solution of the optimization problem (4) vs. the parameter *ϵ*. (B) The optimal cost *J* ^alt^*\** obtained as the solution of the optimization problem (7) vs. the parameter *ϵ*. (C) The optimal cost *J* * obtained as the solution of the optimization problem vs. the control horizon (*t*_*f*_ − *t*_*i*_). (D) The optimal cost *J* ^alt^*\** obtained as the solution of the optimization problem (7) vs. the control horizon (*t*_*f*_ − *t*_*i*_). The parameter *c*_*q*_ and *c*_*p*_ are both set to 1.

From Fig. 4 (C) and (D) we also see the effects of changing the control horizon (*t*_*f*_ − *t*_*i*_) of the optimal control problem, from a minimum of 60 days to a maximum of 120 days. We see here that different cities behave differently. For Houston and LA we see that a longer *t*_*f*_ corresponds to a lower value of the optimal cost *J* *, while for NYC the cost increases with *t*_*f*_. A complete study of the effects of varying *t*_*f*_ can be found in the Supplementary Note 4. In particular, we see that increasing *t*_*f*_ has two contrasting effects on the objective function. On the one hand the cost is integrated over a longer time period, on the other hand the longer time period can be exploited to allow for less stringent measures of social distancing at any time (larger values of *P* *), which may result in a lower value for the objective function overall. Thus it appears that finely tuning the time horizon of the objective function may be used to critically and selectively affect different cities. The implications are particularly significant for those cities, Houston and LA, that seem to need a longer time period to suppress the epidemics. For Seattle we see that *J* * is minimized for intermediate values of *t*_*f*_ in the interval [255, 285], indicating a specific advantage of choosing such a control horizon, see also Fig. 5. A remarkable observation is that by comparing the left plots and the right plots in Fig. 4, it is evident that the optimal control solution is quite independent of the particular form of the objective function (either *J* or *J*^*alt*^.) The reason for this is that when the integrand in (4) is nonlinear the optimal solution maintains *P* *(*t*) as close as possible to the linear regime (high values of *P* *), which is why we do not see much difference with the linear case (7).

**Figure 5:**
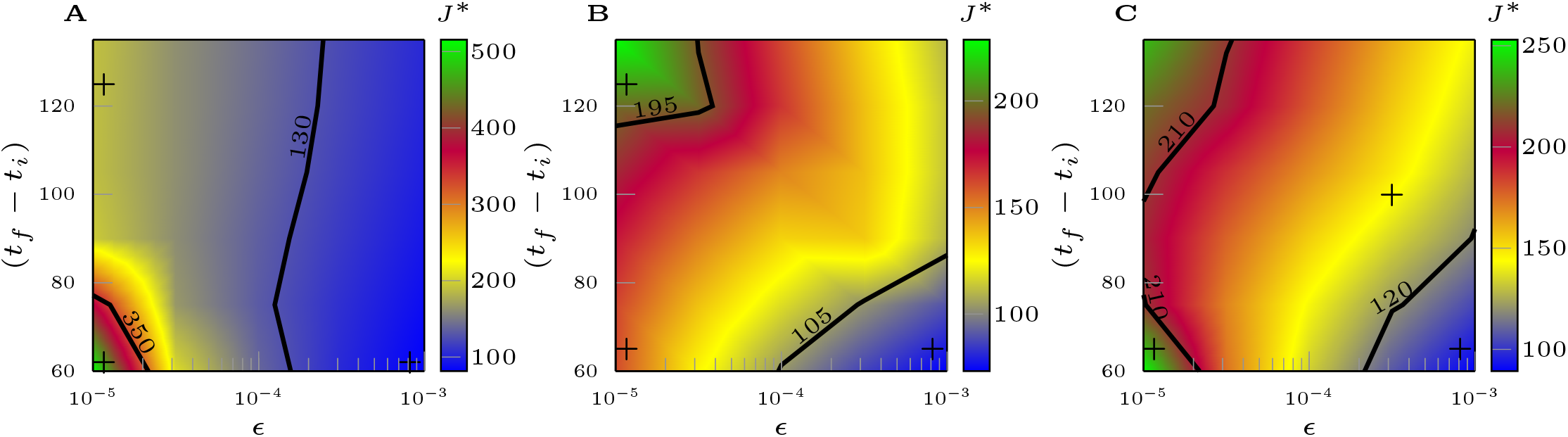
The optimal cost *J* * in the (*t*_*f*_ −*t*_*i*_), *ϵ* plane. (A) shows the case of the Los Angeles Metropolitan Statistical Area. (B) shows the case of the NYC Metropolitan Statistical Area. (C) shows the case of the Seattle Metropolitan Statistical Area. For each city, *I*_max_ is chosen as the maximum of the range in Table 3. Type 1 solutions (in green) are more expensive than type 2 solutions (in blue.) The regions in yellow/red correspond to the transition between the two types of solutions. The parameter *c*_*p*_ and *c*_*q*_ are both set to 1.

In general, the main constraints of the problem are provided by *I*_max_ and *ϵ*. However, for each case considered, typically either one of these two constraints is dominant. In all the simulations shown in Figs. 3, the dominant constraint is provided by *ϵ*, with *I*_*s*_(*t*) + *I*_*tp*_(*t*) remaining well below *I*_*max*_ over the entire period [*t*_*i*_, *t*_*f*_]. These solutions, which we will refer to as the type-1 solution, are characterized by strong measures of social distancing (low *P* *) throughout the whole time interval considered, and the dominant constraint is to achieve suppression of the epidemics at the prescribed terminal time (*t*_*f*_).

We have also seen the emergence of different solutions, which we refer to as the type-2 solution, when the dominant constraint is given by *I*_max_. In these solutions there is at least one time *t* at which the constraint (6) is satisfied with the equal sign. Overall, type 2 solutions are less expensive economic-wise than type 1 solutions, i.e., the value of *J* * is lower. There are also cases when the optimal solution is actively affected by both constraints. In order to better understand the transition between type 1 and type 2 solutions, we have investigated the optimal control problem (4), for the cases of LA, NYC, and Seattle, as both the suppression parameter *ϵ* and the control horizon (*t*_*f*_ − *t*_*i*_) are varied. The transition is characterized by a gradual change in *J* *, high for type 1 solutions (in green) and low for type 2 solutions (in blue), with a transition area in between shown in yellow and red. This is illustrated in Fig. 5 and in more detail in Fig. 19 (LA), Fig. 20 (Seattle) and Fig. 21 (NYC) of the Supplementary Note 5. Our results show that considerations about the timescale of the control action apply differently to different cities. As can be seen, the particular emergence of type-1 or type-2 solutions is affected by both the choice of *ϵ* and (*t*_*f*_ − *t*_*i*_). From Fig. 5 (A) for LA we see that the most expensive control solutions are achieved when one is trying to suppress the epidemics to a low level in a short time (small *ϵ* and short *t*_*f*_ − *t*_*i*_.)

This is opposite to what seen for NYC in Fig. 5 (B), where longer time horizons are usually associated with more expensive solutions. The most convenient solution arises when the suppression constant *ϵ* is large but the control horizon is short (area shown in blue.) Finally, Fig. 5 (C) for Seattle shows that in the case of small *ϵ*, expensive solutions are obtained when the control horizon is either short or long, while an intermediate control horizon is to be preferred. We also see that setting the suppression constraint to a larger value and the control horizon to be short can reduce the cost considerably (which is similar to NYC). In general, these results point out the importance of carefully choosing the timescale over which one is seeking to suppress the epidemics, as well as the suppression level. Intuitively, our results indicate that suppressing the epidemics in a short time is more costly when the epidemics is on the rise (LA) compared to cases when it is not (NYC.)

Next, we briefly consider the case that the transmissibility varies with time, i.e., we replace *β* → *β*(*t*) in Eq. (2). This is consistent e.g., with a situation in which the weather gets colder (such as during the fall season in the Northern emisphere), which has been associated with increased numbers of contagions. We then solve the optimal control problem (2) (4) (5) (6) by computing the optimal control *P* *(*t*) that minimizes the objective function *J*.

The results of our calculations are shown in Fig. 6, where we focus on the New York City Metropolitan Statistical Area and compare the two cases of constant transmissibility (previously studied) and time-varying transmissibility. For simplicity we take *β*(*t*) to increase linearly in time, from the initial value in Table 1 *β*(*t*_*i*_) = 1.806 to the final value *β*(*t*_*f*_) = 2 × *β*(*t*_*i*_) = 3.612. As can be seen, the optimal control solution *P* *(*t*) for the case of linearly increasing *β* differs substantially from the case of time-invariant *β*: in the central phase *P* *(*t*) is now seen to decrease linearly in time, indicating that to a linear increase in transmissibility corresponds a linear decrease in the control parameter (an thus increasingly stricter measures of social distancing). This can be used to plan long-term control interventions over periods of time over which the environmental conditions are subject to well characterized variations that affect the rate of transmissibility of the disease.

**Figure 6:**
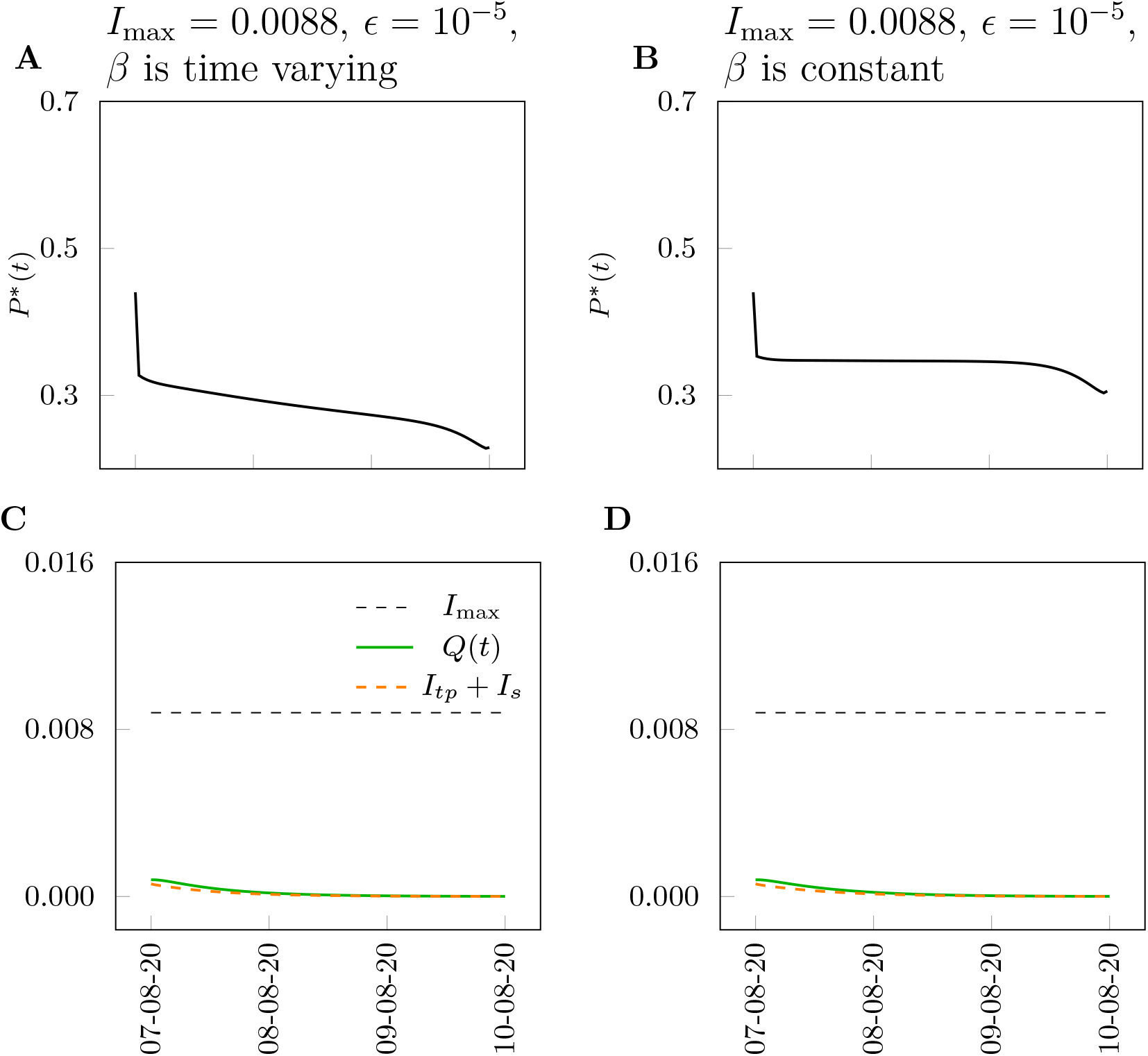
Time-varying transmissibility. We consider the NYC Metropolitan Statistical Area. Left plots: time-invariant transmissibility. Right plots: the transmissibility increases linearly in time. As can be seen, with the exception of the initial phase and of the final phase, the case of linearly increasing transmissibility corresponds to a linearly decreasing optimal control input *P* *(*t*).

Finally, we include here computation of the optimal control solution for a model modified to incorporate the effects of the administration of an effective vaccine against COVID-19. When we initially started working on this paper, there was no certainty about the development and future availability of a vaccine. However, as of February 2021, we are now seeing several effective vaccines being developed, manufactured, and administered in different countries. An important observation is that the production and distribution of the vaccine is a process that takes time. For example, the United States is aiming to administer one million vaccine doses per day [46], which considering the entire US population, corresponds to a time-scale of roughly one year to achieve complete immunization. An important question is thus how the introduction of the vaccine is going to modify the optimal control solutions. While this is not the main focus of this paper, we felt we had to incorporate some preliminary results on the effects of ongoing vaccinations on the optimal control solutions. To be able to provide an answer to this question, we had to (i) modify the model to include the effects of the vaccine being administered to the population, (ii) re-parametrize the model from real data and (iii) compute new optimal control solutions. We briefly outline this process below.

We modeled the effects of administration of the vaccine as an additional flow from the *S* compartment to the *R* compartment. To accommodate for this, we keep using the set of equations (2), but replace Eq. (2a) with the following one,

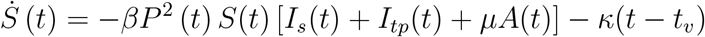

and Eq. (2g) with

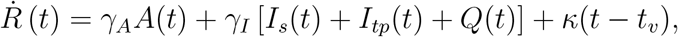

where we take *t* ≥ *t*_*v*_ and *t*_*v*_ is the time the vaccine administration started, which for the United States is 14-Dec-2020. Note that the model accounts for a rate of immunization linearly increasing with time, which was consistent with available data.

In our calculations we focused on the MSA of Seattle, for which we estimated *κ* = 0.000045 (see the Supplementary Information Note 11 for more details.) We then used the same approach outlined before to parametrize the model. The parameters were inferred by the daily case reports over the extended period of time from 21-Jan-2020 to 14-Dec-2020. More information about the parametrization can also be found in the Supplementary Note 11. We then re-computed the optimal control solution by selecting the time at which we start optimizing the control action *t*_*i*_ = *t*_*v*_ and by varying the control horizon *T*_*cont*_ = (*t*_*f*_ − *t*_*i*_). As we already pointed out before, we found the choice of the control horizon to strongly affect the results of the optimization.

Figure 7 shows the results of our computations for the city of Seattle by choosing two different values of the control horizon, *T*_*cont*_ = (*t*_*f*_ — *t*_*i*_) = 130 days (on the left) and *T*_*cont*_ = (*t*_*f*_ − *t*_*i*_) = 150 days (on the right.) In both plots, we compared the two cases that the model includes the effects of vaccination (gray curve, *κ* = 0.000045) or does not (black curve, *κ* = 0.) The full black dot indicates the point in time at which the two curves depart from each other. Both plots show the optimal control solution to deviate at an early time, roughly 15 to 25 days after the beginning of the administration of the vaccine (which corresponds to approximately 6% to 8% of the population getting vaccinated.) This indicates that vaccinations can affect the optimal control solution, even when a relatively little percentage of the population is immunized. In all the simulations we have run we have always seen the optimal control solution to deviate soon after the beginning of vaccinations. Moreover, the separation between the two curves appears to be larger over the longer control horizon. The plot on the right shows the parameter *P* *(*t*) approach 1 at the end of the control horizon, which corresponds to complete removal of the social distancing measures.

**Figure 7:**
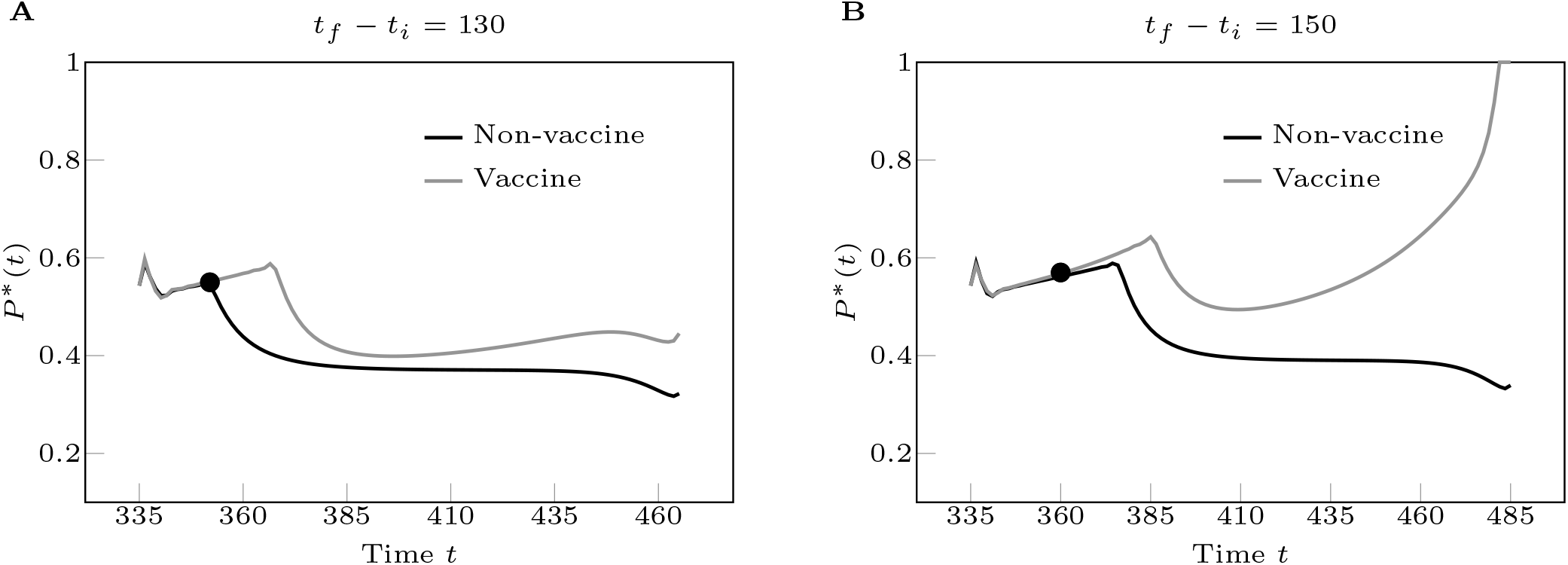
Effects of vaccinations on the optimal control solution. We consider the Seattle Metropolitan Statistical Area. In A the control horizon *T*_*cont*_ = (*t*_*f*_ − *t*_*i*_) = 130 days and in B the control horizon *T*_*cont*_ = (*t*_*f*_ − *t*_*i*_) = 150 days. For both figures, we plot *P* *(*t*) by comparing the two cases that the model includes the effects of vaccination (gray curve) or does not (black curve.) The full black dot indicates the point in time at which the two curves depart from each other. As can be seen the separation between the two curves appears to be much larger over a longer control horizon. The plot on the right shows the parameter *P* *(*t*) approach 1 at the end of the control horizon, which corresponds to removal of the social distancing measures. Additional plots showing the time evolutions of the states corresponding to the optimal solutions in A and B can be found in the Supplementary Information, Note 11.

Our results point out that in a realistic scenario the optimal level of social distancing is affected by the introduction of vaccinations, especially over longer periods of time. This is not surprising as the expectation is that the long-term effect of administration of the vaccines will be relaxation of the social distancing measures. However, the advantage of our analysis is that it can be used to assess the most economically advantageous level of social distancing *while the vaccines are being administered to the population*. For example, Fig. 7B shows substantial relaxation of social distancing roughly five months after the date in which administration of the vaccines began. We remark that the derived timescale (150 days) is subject to the simple vaccination model, in which we optimistically assume that (1) all the doses are reserved for the susceptible only, and (2) the production of the vaccines remains linear in this 150 days. Our simulation shows that, with these two assumptions, at the end of this period herd immunity is achieved and social distancing is removed. However, such a timescale can be treated as an optimistic forecast; the real timescale to reach herd immunity could be longer due to the violation of both of the assumptions. It is important to point out once again that the optimization we perform is one for which the goal is to minimize the impact of social distancing on the economy, in the presence of relevant public health constraints.

## Discussion

Our analysis reveals that the cost of eradicating the disease—i.e., suppressing the number of infected individuals down to a certain critical threshold, such as « 1 person in the formulation of our stylized mathematical model, even in the optimal case, is significantly higher than the cost of ‘managing’ the pandemic to avoid the saturation of regional medical resources. In light of the current progression of the pandemic in the US, our analysis brings rigorous scientific and quantitative foundation for the latter strategy, which is adopted by many local administrations (e.g., State offices.)

Our conclusion that the optimal control solution is well approximated by a constant level of social distancing contrasts with the implemented system of reopening ‘in phases’ [14]. In many countries, the expected progression is from phase 1 to phase 2 and higher, but there have been several cases in which a premature reopening followed by a rise in COVID-19 numbers has led to folding back into phase 1 [41, 44, 17, 22]. Based on our results, such alternating control interventions are suboptimal because the economic benefits of momentarily relaxing the restrictions are lower than the costs associated with the successive tightening (such as e.g., a second lockdown.) Instead we have shown that in the presence of smoothly time-varying environmental conditions (e.g., affecting the transmissibility), the optimal control solution also varies smoothly in time. This allows us to plan long-term control interventions over periods of time over which the environmental conditions are subject to well characterized variations that affect the rate of transmissibility of the disease.

Several countries have adopted control strategies that are region-specific. For example, Italy has assigned to its regions color-coded restrictions (yellow, orange, and red) [31], which, based on a careful monitoring of the local progression of the disease, have been updated frequently. Our model focuses on individual MSA’s (or regions) and as such neglects the larger-scale spatial resolution of the epidemic. The need for frequent updates of the control interventions provides evidence of the inherent difficulty of ‘closing the control loop’, due to many possible factors, such as partial and incorrect measurements, delays in obtaining information and implementing actions, imperfect application of the controls, lack of resources, the effects of people traveling from area to area, and others. While all these factors would require separate consideration, our main recommendation that interventions should be maintained at a (nearly) constant level stands. Our work indicates that the best intervention for the economy is one that does not fluctuate in time, while alternating control actions are suboptimal. In addition, frequently changing interventions present other disadvantages which have made them unpopular [8]: they are difficult to implement, and to follow, and they negatively affect business planning capability.

Occasionally, there have been claims by policy makers and/or scientists that seeking herd immunity by exposure to the virus (in the absence of a vaccine) may provide the best long-term path-forward [30, 20]. However, the timescales over which herd immunity can be achieved without violating the *I*_max_ constraint appear to be quite long (see Methods) in the absence of a vaccine. By setting a very large control horizon in our simulations, we have seen the emergence of these ‘herd immunity solutions’, shown in the Supplementary Note 6. However, the parameters of our realistic scenarios, estimated from real data, are far away from the point at which such solutions are optimal. In general, our study allows to quantify how ‘far’ one is in the parameter space from the point at which a herd immunity solution becomes optimal.

## Conclusion

In this paper, we have proposed an approach to optimize social distancing measures in time in order to contain the spread of the COVID-19 epidemics, while minimizing the impact on the economy. Our analysis has been applied to four different metropolitan statistical areas within the US, but can be directly applied to other geographical areas. Each MSA was shown to be well described by a stylized mathematical model whose parameters were inferred by daily new case counts reported by local health administrations. Our approach based on modeling, inferring model parameters from real data, and computing optimal control solutions for the inferred model is general and may be applied to other complex systems of interest.

Our choice of the objective function is quite simplified, namely we assume that increasing levels of social distancing and quarantining result in progressively higher economical costs. In addition, we did not account for the cost associated with medical treatments, which is important yet significantly smaller than the cost of shutting down the economical activities of the entire society [10]. Our approach can be easily generalized to other more complex, more realistic types of objective functions, see e.g., [42, 43]. Different from this study, these objective functions may also be specific to given regions or try to capture a particular socio-economical model of interest.

We have found that the optimal control solutions are quite robust to the specific choice of the objective function and of its parameters, such as *c*_*q*_. These control solutions tend to be qualitatively similar for different cities. However, they are affected by the choice of the constraints, in particular the constraint that we have associated to suppression of the epidemic, and by the time horizon of the control action. When these are varied, different cities behave differently, which points out the importance of our data-driven approach. We have also seen that small deviations from the optimal solution can lead to dramatic violations of the constraints.

It is possible to translate these optimal interventions in actual measures that can be imposed on the population, such as restricting the access to certain businesses or venues. Available technology includes the usage of coupled digital non-contact healthcare systems [35]. While implementation of a time-varying control may be challenging in practice, we found that the optimal solution is typically characterized by an initial drop (due essentially to the non-optimality of current control interventions), followed by a nearly constant control (specific to each city) for a long time, and by a final drop, which is needed to achieve the desired suppression of the epidemics at the final time. Thus the optimal control solution is almost constant except for the initial time and the final time, which significantly increases the applicability of this study. The constant part of the solution could be practically achieved by dynamical regulation to control the time-varying reproduction number *R*_*t*_ ≈ 1. A key observation is that the optimal solution *P* * is generally lower than the value of *P* inferred by data. The initial drop in *P* *\ may provide a measure of non-optimality of current interventions. This drop was seen to be smallest for NYC compared to other US cities, but was present in all the cities we have analyzed.

The time-scale of control interventions, i.e., the control horizon, appears to play a fundamental role. Considerations about the control horizon may vary from area to area and may be affected by a number of factors, including the times at which a vaccine becomes available and is distributed to the population. It appears that cities that have seen an increase of cases during the inference period need a longer control horizon to suppress the epidemics optimally. In certain instances, the impact on the economy can be minimized by tuning the control horizon; for example, for the city of Seattle we found that an optimal control horizon was equal to roughly 90 days when the suppression constraint *ϵ* = 10^−5^. We wish to emphasize that given the very large economical impact of social distancing measures, even a small improvement in the control strategy can lead to considerable economical benefits.

We also computed the most economically advantageous level of social distancing during the time over which the vaccines are administered to the population. Our simulations show that the optimal control solution is affected by the introduction of the vaccine quite early on, which indicates the possibility of gradually relaxing measures of social distancing soon after the beginning of vaccinations and strongly reduce them roughly five months later. We acknowledge that our study on the effects of the vaccines is limited, as we only focus on one region and we see that the optimal control solution is strongly affected by the choice of the control horizon. We also do not consider the emergence of new variants against which existing vaccines may be less effective. Our assumption of a linearly increasing rate of vaccination of the population should be validated by using region-specific data. Extending the analysis to other regions and incorporating more realistic data would require a major effort that is beyond the scope of this paper. Our conclusion that over longer time periods vaccination would allow substantial reductions in the level of social distancing, should be taken with the due precautions. One should be reminded that our objective in this paper is to minimize the impact of social distancing on the economy, in the presence of relevant public health constraints. Nonetheless, our approach based on modeling, model parametrization, and optimal control, could be easily adapted to different geographical areas to provide region-specific recommendations.

Other limitations of our work are briefly discussed below. Limited testing capabilities may affect some of our results. The model could benefit from incorporation of considerations about spatial effects. Many regulations have been introduced to limit people’s mobility during the pandemic; however essential travel has often remained in place, which has probably been responsible for much of the disease’s propagation. One first step to incorporate spatial resolution in the model would be considering an extension of the model with two communicating regions. This could be the subject of future work.

One relevant question is whether policy-makers can assess whether a currently employed control action is optimal or not. The so-called HAMVET procedure, initially proposed in [38] and presented in the Supplementary Note 9, can be used to validate a control strategy and evaluate its optimality.

## Data Availability

Data was acquired from the New York Times GitHub repository https://github.com/nytimes/covid-19-data

## Methods

### Estimation of *I*_max_

The *I*_max_ values can be approximated with simple assumptions as seen in Table 3 for a selection of major U.S. cities. These *I*_max_ values are a ratio of ICU beds to the population of people which require them. Following [23], we estimate that the probability of death conditioned on symptomatic infection is equal to *f*_*H*_ ×(1−*f*_*R*_) = 0.01134, where the two parameters *f*_*H*_ and *f*_*R*_ were independently computed in [32] and [37], respectively. Data [33, 16] shows that the mortality rate for patients sent to ICU is between 30% and 40%, thus, it is reasonable to assume that an overall fraction of infected people equal to 0.01134*/*0.35 = 0.0324 needs ICU beds. In the hypothetical situation that the population of an entire state contracts COVID-19 3.24% will require an ICU bed. The *ρ* term is a modifier which denotes how many ICU beds are available to COVID-19 patients as some beds could be used for other reasons. Reasonably, the value of *ρ* ranges from 2/3 to 1 in Table 3. *I*_max_ is then calculated as the number of available ICU beds (including the *ρ* assumption) divided by number people which contract COVID-19 and also require an ICU bed (3.24% assumption).

For all our numerical experiments, we consider two values for *I*_max_, one corresponding to *ρ* = 2*/*3 and another one corresponding to *ρ* = 1.

### Timescale for *T*_herd_

We now attempt to answer the following question. By enforcing satisfaction of the constraint with the equal sign *I*_*s*_(*t*) + *I*_*tp*_(*t*) = *I*_max_, how long would it take before herd immunity is achieved? By assuming long-term immunity of those recovered from the virus, we can expect herd immunity to arise when roughly 80% of the population has been exposed [36]. Consider for example the case of NYC for which from Table 3 we see that *I*_max_ is between 0.88 % and 1.32 %. We assume an average hospitalization of 20 days [25]. That means that the time to achieve herd immunity *T*_herd_ varies between 606 days=(80 ×20*/*(1.32 × 2)) and 909 days=(80 × 12*/*(0.88 × 2)). The factor of 2 accounts for the fact that roughly one exposed person out of two develops symptoms. Analogously, for Chicago, we estimate *T*_herd_ to vary between 533 days=(80 × 20*/*(1.5 × 2)) and 800 days=(80 × 20*/*(1 × 2)). From these back-of-the-envelope calculations we see that the timescale over which herd immunity can be achieved without violating the *I*_max_ constraint appears to be quite long and comparable to if not longer than the timescale over which we expect availability of a vaccine.

## Acknowledgements

The authors would like to thank Bill Hlavacek, Isaac Klickstein, Neethi Thevan T., and Enrico Del Frate for insightful discussions. YTL thanks the support from the Laboratory Directed Research and Development Program at Los Alamos National Laboratory (Project XX01).

